# Epigenome-wide association study of posttraumatic stress disorder in a South African population

**DOI:** 10.1101/2025.05.07.25327159

**Authors:** Sylvanus Toikumo, Seyma Katrinli, Morné Du Plessis, Stefanie Malan-Müller, Leigh L. van den Heuvel, Gerard Tromp, Alicia K. Smith, Soraya Seedat, Sian MJ Hemmings

## Abstract

Posttraumatic stress disorder (PTSD) is a disabling psychiatric disorder with both genetic and environmental contributions. Epigenetic mechanisms have been found to, at least partially, mediate the effect of environmental factors in PTSD aetiology. PTSD has also been found to be associated with all-cause morbidity and premature mortality, suggesting accelerated biological aging. However, the role of PTSD-associated epigenetic changes and aging in sub-Saharan African subjects is yet to be elucidated. We address this knowledge gap by conducting an epigenome-wide association study (EWAS) of clinician-diagnosed PTSD in 120 individuals from a 5-way genetically admixed South African population (PTSD: n=61; trauma-exposed controls: n=59). Genome-wide DNA methylation (DNAm) data was generated using the Illumina MethylationEPIC BeadChip (V1). EWAS analysis was conducted using a mixed-effects regression model adjusted for sex, age, cell-type proportions, principal components (PCs), and positional effects. Epigenetic age acceleration was calculated using Horvath, Hannum, PhenoAge, and GrimAge clocks. Three less methylated CpG sites and thirteen differentially methylated regions (DMRs) were associated with PTSD. PTSD severity, but not PTSD diagnosis, was associated with accelerated Horvath DNAm age (*p* = 0.039). Furthermore, individuals with PTSD and comorbid metabolic syndrome (MetS) exhibited significantly accelerated DNAm age compared to those without MetS, as measured by the Hannum (*p* = 0.027), Horvath (*p* = 6.22 × 10⁻⁴), and Levine PhenoAge (*p* = 0.039) clocks. These findings suggest an increased risk of premature mortality in individuals with PTSD and comorbid MetS. This study highlights novel PTSD-associated epigenetic differences within a South African population and offers new insights into the role of age-related epigenetic mechanisms in PTSD susceptibility and progression.

## Introduction

Posttraumatic stress disorder (PTSD) is a debilitating disorder that leads to substantial impairment or distress^1^. Approximately 75% of the South African population encounter at least one traumatic event throughout their lifetime, while more than half will experience multiple traumatic events^2–4^. Estimates placing the prevalence of PTSD in South Africa (SA) at approximately 2-3% are likely not truly reflective of the general population, since mental health stigmatization and poor retrospective recalling of traumatic events likely contribute to historical underreporting and underdiagnosis^4,5^.

Gene-environment interactions may contribute to the risk of developing PTSD^6,7^. Epigenetic processes provide a mechanism through which gene-environment interactions can affect disease outcomes^8^. Epigenome-wide association studies (EWAS) of PTSD have identified differential DNA methylation (DNAm) profiles within genes involved in hypothalamic-pituitary-adrenal (HPA) axis regulation, central nervous system (CNS) development, synaptic plasticity, and immune system regulation^9–17^. Whereas most EWAS in PTSD have been conducted in predominantly European populations^9–13,15,16^, studies conducted in African populations have focused on women^14^ or combat-related samples^17,18^. Identifying PTSD-related DNAm differences in individuals outside these demographic paradigms is critical for ensuring translational research equity for the general global population.

Trauma burden and PTSD are also associated with poor health outcomes and mortality^19^. This association may be partly due to the increased prevalence of age-related diseases (such as diabetes and cardiovascular disease [CVD]) in individuals with PTSD^20^. It is hypothesized that the increased incidence of such age-related conditions is due to premature aging processes, i.e., an accelerated rate of cellular aging. Of particular interest is the association between PTSD and metabolic syndrome (MetS)^19^, a cluster of traits (including abdominal obesity, high blood pressure and plasma glucose, and dyslipidemia) indicative of an increased risk for CVD^21^. Moreover, individuals in sub-Saharan Africa (SSA) are at greater risk for MetS than those in the global North, significantly increasing the burden of disease and healthcare costs^22,23^. To develop effective treatments for PTSD, it is therefore imperative to identify modifiable molecular biomarkers associated with its etiology and its comorbidity with MetS.

DNAm profiles at specific genomic sites are linked to chronological age and can serve as biomarkers for inferring accelerated cellular aging, regardless of tissue type^24,25^. Various DNAm clocks, like Hannum^24^, Horvath^26^, Levine^27^, and GrimAge^28^, are used to measure biological age in disease progression^25^, but studies on PTSD have shown mixed results, with some indicating accelerated^29^ or decelerated^30,31^ DNAm aging and others showing no significant differences^29,32,33^. Variable findings may stem from differences in prediction models^34^ or trauma factors^35^, while MetS has consistently shown accelerated aging using both the Hannum and Horvath clocks^36–38^. The GrimAge epigenetic clock is a strong predictor of age-related health outcomes^28^ and has been linked to PTSD diagnosis^39^, symptom severity^40^, and cortical thickness in emotion-related brain areas^39^. It is also associated with MetS, especially in European populations^41,42^, with accelerated GrimAge being more common in veterans with both PTSD and obesity^43^. However, no studies have explored PTSD-MetS comorbidity in sub-Saharan African populations, raising the question of whether GrimAge patterns differ between continental and diaspora Africans.

In the present study, our primary aim was to identify genome-wide DNAm profiles associated with PTSD in a South African population. As a secondary aim, we investigated the association between DNAm age and PTSD, and its comorbidity with MetS in the same cohort.

## Methods

### Participants

The current study is nested within a larger initiative, known as “Shared Roots”, the aim of which is to investigate the shared molecular underpinnings of neuropsychiatric disorders (PTSD, Parkinson’s disease, and schizophrenia) and CVD risk^44^. Stellenbosch University Health Research Ethics Committee approved this study prior to data collection (Ethics approval number: N13/08/115), and written informed consent was obtained from all participants.

Study participants were recruited through purposive sampling: (i) from general and psychiatric hospitals and community clinics within the catchment area around Tygerberg hospital; (ii) via print, radio and web advertisements; and (iii) referral from the Mental Health Information Centre in the Department of Psychiatry, Faculty of Medicine and Health Sciences, Tygerberg. Participants in the study were from the South African mixed ancestry/South African Coloured (SAC) population. The SAC population, from which participants were recruited, is a uniquely South African genetically admixed population^45^ derived from five ethnicities (Khoesan, Bantu-speaking Africans, European, and East and Southeast Asian)^46^. They are one of the main population groups in the Cape Town area and form more than 50% of the total population of the Western Cape Province^47^. Adult patients aged ≥ 18 years who self-identified as SAC and with a record of trauma exposure, with/without PTSD were included in this study. All participants had to be able to read and write in English or Afrikaans. The exclusion criteria were adults with any neurological disorder, or an alcohol or drug use disorder within the past 12 months, due to potential confounding.

### Clinical assessment

All clinical assessments were administered by clinicians and trained research psychologists within the Department of Psychiatry, Stellenbosch University. Trauma exposure was assessed with the 16-item self-report Life Events Checklist for DSM-5 (LEC-5)^48^. The Diagnostic and Statistical Manual of Mental Disorders (DSM–5) definition of trauma-exposure was employed^49^. PTSD caseness was defined as meeting the DSM-5 criteria of PTSD according to the diagnostic evaluation with the Clinician Administered Post-traumatic Stress Disorder Scale for DSM–5 (CAPS-5)^50^. Trauma exposed participants in the PTSD cohort not meeting the DSM-5 criteria on the CAPS-5 were designated as trauma-exposed controls (TEC).

The harmonized Joint Interim Statement was used as a diagnostic measure for metabolic syndrome (MetS)^51^. The diagnosis of MetS required the presence of at least three of the following five risk factors: (i) elevated blood pressure (systolic ≥ 130 or diastolic ≥ 85 mmHg) or antihypertensive treatment; (ii) elevated triglycerides (> 1.70 mmol/l or 150 mg/dl) or treatment for hypertriglyceridemia; (iii) low HDL-C (< 1.0 mmol/l for males, < 1.3 mmol/l for females) or treatment for low HDL-C; (iv) elevated fasting glucose (≥ 5.6 mmol/l or 100 mg/dl) or treatment for diabetes; (v) elevated waist circumference (≥ 90 cm for both males and females)^52^.

### Biological data

Approximately 10-15 ml of blood was drawn from all participants (N = 120) and collected in three 5 ml ethylene-diamine-tetra-acetic acid tubes. DNA extraction from 3 ml of whole blood was performed using the Gentra Puregene Blood kit according to the manufacturer’s instructions^53^. The concentration of DNA was determined using an ultraviolet-visible (UV-Vis) spectrometer [Nano-Drop 2000 (ThermoFisher)]^54^, and each sample was electrophoresed on a 1% agarose gel, ensuring that only high quality, high molecular weight DNA were selected for methylation analyses. DNA samples were quantified by fluorimetry [Qubit fluorometer (ThermoFisher)]^55^.

### Genome-wide DNA methylation

Genome-wide methylation analysis, including bisulfite treatment, was performed at the University of Southern California Molecular Genomics Core Facility (USA) (https://uscnorriscancer.usc.edu/core/molgen/). Approximately 1 μg of DNA was subjected to bisulfite treatment using the EZ DNA Methylation Lightning Kit (Zymo Research)^56^ to convert all unmethylated cytosine residues to uracil and retain all methylated cytosines within the samples. Controls for DNA methylation consisted of a total methylated control (enzymatically methylated Jurkat genomic DNA), an intermediate-methylated control (genomic Jurkat DNA) and non-methylated control (whole-genome amplified genomic DNA). To determine the efficiency of bisulfite conversion, the bisulfite treatment was performed on technical controls within the same experiment.

DNA methylation levels were interrogated using the MethylationEPIC BeadChip (Illumina)^57^. This platform offers comprehensive genome-wide methylation profiling at single-nucleotide resolution, targeting sites within promoter regions, open chromatin and enhancer regions, DNase hypersensitive sites, miRNA promoter regions, 5’-untranslated regions (UTRs), first exons, gene bodies and 3’-UTRs^57^. The Illumina Infinium HD Assay for methylation was used to perform whole-genome methylation according to the manufacturer’s protocol^57^. The level of methylation at each locus was determined using the HiScan scanner (Illumina)^57^.

### Quality control (QC)

We used the R package *ewastools* (v1.7.2)^58^ to evaluate 17 control metrics which are described in the BeadArray Controls Reporter Software Guide from Illumina^57^, and as previously described^59^. No samples failed the control metrics. *CpGassoc*^60^ was used to filter out samples with probe detection call rates <90% and an average intensity value of either <50% of the experiment-wide sample mean or <2000 arbitrary units (AU). Low-quality probes with detection p-values >0.01 were set to “missing” and were filtered out for >10% of samples within studies. We removed cross-hybridizing probes^61^.

For each sample, cellular heterogeneity (i.e., the proportion of CD8+ T, CD4+ T, natural killer (NK), B cells, monocytes and neutrophils) was predicted using the Robust Partial Correlation (RPC) method implemented in *Epidish*^62^ using the reference data reported by Salas et al^63^. Ancestry principal components (PCs) were generated from DNAm, following the method described by Barfield et al.^64^, as previously implemented^65^. The components that correlated most with self-reported race/ethnicity (PCs 2-3) were used to adjust for ancestry^64,65^.

### EWAS and DMR analyses

We accounted for chip batch effects by fitting a random intercept model that included chip as a random effect, using R package *CpGassoc*^60^, as published previously^59^. Logit transformed ß-values (M-values) were modeled by mixed-effect regression as a function of PTSD, adjusting for sex, age, CD8+ T, CD4+ T, NK, B cell, and monocyte cell proportions, and ancestry using PCs, and including chip as a random effect term. The R package *BACON* (v3.21)^66^ was used to adjust for inflation of the t-statistics and p-values. The EWAS threshold of 3.6 × 10^-8^ was used to identify significant CpGs^67^. Post hoc sensitivity analysis explored the possible confounding effects of smoking by including DNAm-derived smoking scores as a covariate.

We also assessed differentially methylated regions (DMRs) using the *dmrff* R package (v1.1.2)^68^. DMRs were defined as regions containing two or more CpG sites spanning ≤500 bp, with EWAS analysis *p* ≤ 0.05 and methylation changes in a consistent direction. Following the subregion selection step in *dmrff*, DMRs with FDR-adjusted *p* ≤ 0.05 were considered significant.

### Secondary/exploratory analyses

For the significant CpG sites or DMRs, we examined potential associations with PTSD-related health outcomes based on existing literature in the EWAS Catalog^69^ and EWAS Atlas^70^. Correlations between blood and brain DNAm levels were examined for EWAS and DMR significant hits using the IMAGE-CpG database, which contains DNAm measures from blood, saliva, buccal, and live brain tissue samples from 27 patients with medically intractable epilepsy undergoing brain resection^71^. We also conducted exploratory over-representation analysis using the “Web-based GEneSeT AnaLysis Toolkit” (WebGestalt) (www.webgestalt.org)^72^. Given the number of EWAS significant hits (*n* = 3) and potential gene-gene correlations below conservative threshold values^73^, we considered CpGs that showed suggestive associations (*p* < 5×10^-4^) for gene-set analysis. Bonferroni correction (*p* < 0.05) was applied to identify gene sets significantly associated with PTSD.

### DNAm age analyses

To determine the association between accelerated DNAm age and PTSD, DNAm age was calculated based on four clocks — Hannum, Horvath, Levine, and GrimAge — using the *dnaMethyAge* R package (v0.2.0)^74^. We tested the correlations between accelerated DNAm age (the residual obtained from regressing each DNAm age clock on chronological age) and potential confounders^75^, including sex, and cellular heterogeneity. Associations of DNAm age acceleration with PTSD diagnosis (model 1), PTSD severity (based on CAPS score) (model 2), and PTSD-MetS comorbidity (using an interaction term; model 3) were evaluated using linear regression models, adjusting for sex, and the estimated proportions of CD8+ T, CD4+ T, NK, B cells and monocytes.

*Model 1: DNAm age ∼ PTSD + sex + cell types*

*Model 2: DNAm age ∼ PTSD severity (CAPS Score) + sex + cell types*

*Model 3: DNAm age ∼ PTSD*MetS + sex + cell types*

## Results

### Demographic and clinical characteristics

Our sample comprised 120 individuals, of whom 59 (49.17%) were trauma-exposed controls (TEC) without any history of PTSD, and the remainder were diagnosed with PTSD. The mean chronological age was 43.2 years (SD = 10.8, range 19.9 – 64.7), with no significant differences observed between PTSD cases and TEC groups (*p* = 0.64). Females represented 73.8% and 67.8% in PTSD and TEC groups, respectively (*p* = 0.48) (Table 1). The mean CAPS score differed significantly between PTSD cases and TECs (36.4 ± 7.5 in cases; 7.1 ± 6.9 in TEC; *p* = 2.51 × 10^-44^). Approximately 48% of PTSD cases and 49% of TECs met the criteria for MetS (Table 1).

**Table 1:** Study demographic and clinical characteristics.

| | PTSD (N = 61) | TEC (N = 59) | $p$ |
| --- | --- | --- | --- |
|  | Mean (SD) or N (%) |  |  |
| Sex, Female | 45 (73.8%) | 40 (67.8%) | 0.48 |
| Age | 42.7 (10.7) | 43.6 (10.9) | 0.64 |
| CAPS score | 36.4 (7.5) | 7.1 (6.9) | $2.51 \times 10^{-44}$ |
| MetS, Present | 29 (47.5%) | 29 (49.2%) | 0.86 |
| Current smoking % | 27 (44.3%) | 22 (37.3%) | 0.44 |
| Hannum DNAm age acceleration | 0.14 (4.0) | -0.15 (4.0) | 0.64 |
| Horvath DNAm age acceleration | 0.54 (4.7) | -0.56 (4.1) | 0.17 |
| Levine PhenoAge acceleration | -0.27 (5.1) | 0.28 (4.6) | 0.53 |
| GrimAge acceleration | -0.20 (3.2) | 0.21 (3.3) | 0.49 |

PTSD – Posttraumatic stress disorder; MetS – Metabolic syndrome; TEC – Trauma exposed control; SD – Standard deviation

### PTSD-associated CpGs and biological pathways

Three CpG sites (cg01603456: intergenic, cg00511884: *LINC00299,* and cg26847571: *SYNM*), were found to be significantly lower in PTSD cases compared to TECs (Table 2, Fig 1A). Of the three PTSD-associated CpGs, two (cg01603456: intergenic and cg00511884: *LINC00299*) remained significant with the same direction of association in the sensitivity analysis that adjusted for smoking score (Table 2), indicating that the PTSD-associated DNAm profiles in these regions were largely unaffected by smoking. cg26847571 (*SYNM*) methylation was not significant after correcting for the effect of current smoking.

**Figure 1.**
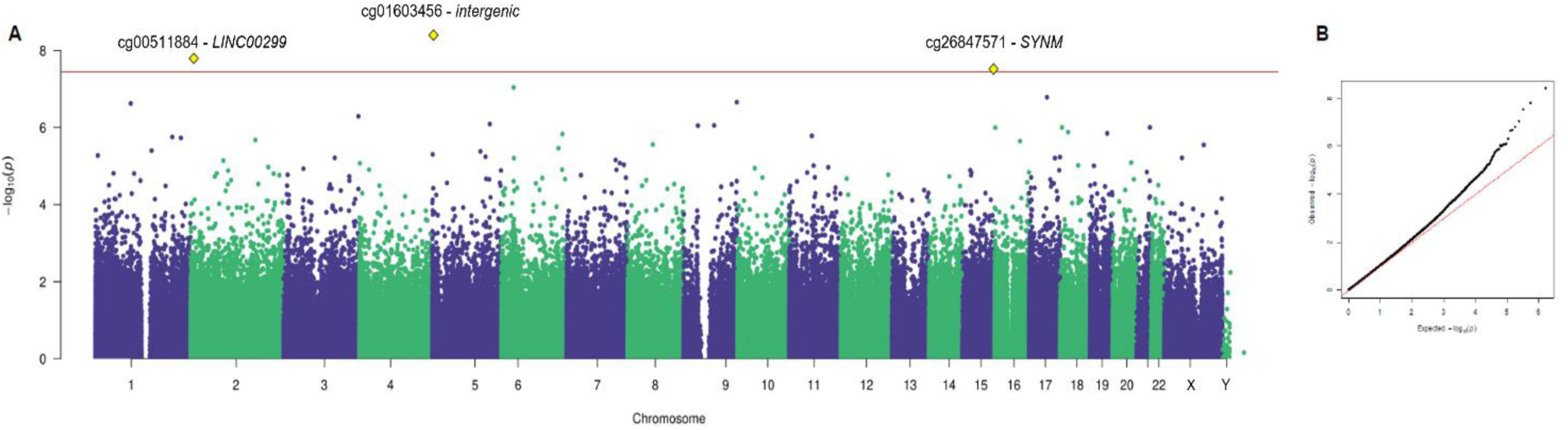
PTSD associated DNA methylation sites across the genome. A. Manhattan plot for epigenome-wide association analysis across 120 individuals. B. Quantile-quantile plot showing no evidence of genomic inflation (λ = 1). The *x*-axis of the Manhattan plot is the chromosomal location of each CpG site, and the *y*-axis is the −log 10 of the unadjusted *p-*value for the association with PTSD. The red line indicates genome-wide EWAS statistical significance at *p* < 3 × 10^-8^.

**Table 2:** Epigenome-wide significant CpG sites associated with PTSD.

| CpG | Location | Gene | Features | Beta | SE | *p* |
| --- | --- | --- | --- | --- | --- | --- |
| *Before sensitivity analysis* |  |  |  |  |  |  |
| cg01603456 | chr5:3324867 | intergenic | Island | -0.16 | 0.03 | 4.01E-09 |
| cg00511884 | chr2:8374459 | LINC00299 | Body; 3'UTR | -0.10 | 0.02 | 1.61E-08 |
| cg26847571 | chr15:99664049 | SYNM | Body; 5'UTR | -0.25 | 0.05 | 3.05E-08 |
| *After sensitivity analysis* |  |  |  |  |  |  |
| cg01603456 | chr5:3324867 | intergenic | Island | -0.16 | 0.03 | 3.94E-09 |
| cg00511884 | chr2:8374459 | LINC00299 | Body; 3'UTR | -0.10 | 0.02 | 1.35E-08 |

Epigenome-wide significant CpG sites for PTSD were identified using the EWAS threshold $p < 3.6\text{E-}08$ . Chr – chromosome, UTR – untranslated region, SYNM - synemin

We also identified 13 DMRs that were significantly associated with PTSD (FDR < 0.05, Supplementary Table 1). CpGs within the DMRs mapped to genic regions that included *AIRE*, *CARKD*, *BDNF-AS*, *MAP3K6*, *RUSC1*, *Cxorf57* and *CUGBP1*, among others (Supplementary Table 1)

Based on data from the IMAGE-CpG database^71^, blood DNAm levels of a total of seven CpG sites (cg01603456, intergenic; cg22877509, cg19516515 in RUN and SH3 domain containing 1 gene [*RUSC1*]; cg19398192, cg14057181, cg11111131 in *CXorf57*; and cg16742756 in CUG triplet repeat, RNA binding protein 1 gene [*CUGBP1*]) were found to correlate with brain DNAm levels (Supplementary Table 2). Using data from the EWAS Catalog^69^ and EWAS Atlas^70^, our results were concordant with EWAS hits for C-reactive protein levels, aging, alcohol consumption, HIV infection, and rheumatoid arthritis, among others (Supplementary Table 3).

Gene-set enrichment analysis identified significantly enriched pathways related to Rap signaling (hsa04015) and dopaminergic synapse (hsa04728) pathways (FDR < 0.05; Supplementary Table 4). Cellular components, including neuron projection (GO:0043005) and synapse (GO:0045202), were also enriched in the PTSD group (FDR < 0.05; Supplementary Table 4).

### Epigenetic age acceleration in PTSD

All four DNAm age clocks (Hannum, Horvath, Levine PhenoAge, and GrimAge) were positively correlated with chronological age, with correlation estimates ranging from 0.87 to 0.93 (*p* < 0.001; Supplementary Figure 1). GrimAge acceleration correlated negatively with B cell proportions (r = -0.25; *p* = 0.04; Supplementary Figure 2) and Levine PhenoAge acceleration negatively correlated with CD4+ T levels (r = -0.33; *p* = 0.01) and B cell levels (r = -0.26; *p* = 0.04) (Supplementary Figure 2). Sex showed no significant correlation with either Hannum or Horvath DNAm age acceleration.

PTSD symptom severity, but not PTSD diagnosis, was significantly associated with accelerated Horvath DNAm age (*β* = 0.05, se = 0.02, *p* = 0.039; Supplementary Table 5). We also observed a significant association between PTSD-MetS comorbidity and accelerated aging based on three of the DNAm clocks, namely Hannum DNAm age (*β* = 1.87, se = 0.83, *p* = 0.027), Horvath DNAm age (*β* = 3.03, se = 0.86, *p* = 6.22 × 10^-4^), and Levine PhenoAge (*β* = 1.96, se = 0.94, *p* = 0.039) (Supplementary Table 5).

## Discussion

This study presents an epigenomic investigation of PTSD in 120 South African mixed ancestry individuals. We profiled DNAm at the genome-wide scale in whole blood and identified three CpG sites and 13 DMRs that were significantly differentially methylated in individuals with PTSD compared to TECs.

Of particular interest in the context of PTSD are the EWAS findings demonstrating decreased methylation of a CpG site (cg00511884) in *LINC00299* in individuals diagnosed with PTSD compared to TECs. *LINC00299* is a non-coding RNA on chromosome 2 and disruption of LINC00299 could play a role in abnormal neurodevelopment^76^. Genetic variation in *LINC00299* has previously been associated with current tobacco smoking^77^, with continued smoking in a longitudinal study, and with metabolic traits such as bilirubin and N-acetylornithine levels^78–80^. In a recent study, decreased methylation at CpG site (cg23079012) in *LINC00299* was associated with cannabis use disorder (CUD) in a veteran cohort enriched for PTSD after controlling for current smoking^81^. In particular, PTSD and CUD interacted to affect cg23079012 such that veterans with both disorders showed significantly lower DNAm levels compared to the other groups. In the current study, the sensitivity analysis demonstrated that lower DNAm levels of cg00511884 in *LINC00299* remained significantly associated with PTSD independent of smoking, suggesting that methylation of this particular CpG site may be specific to PTSD. Further investigations are warranted to disentangle potential pleiotropic effects of DNAm changes in cg23079012 on comorbid psychiatric conditions, such as CUD, and to better understand the role that metabolic traits may play in these associations.

Our results also demonstrate an association between a hypomethylated site in the *SYNM* gene body and PTSD. Earlier findings revealed increased expression of *SYNM* in peripheral blood mononuclear cells from trauma survivors with PTSD, suggesting a role in stress response^82^. Differential methylation and expression of *SYNM* could be a mechanism that explains stress-related biological responses in individuals with PTSD. SYNM is involved in mitochondrial metabolism, and DNAm of a CpG site located near *SYNM* (cg16765088) has been found to be associated with T2D in a European population^83^. Moreover, altered SYNM protein levels in individuals with atrial fibrillation have been linked with mitochondrial metabolic changes of the atrium^84^. More research is needed to shed light on the role of DNAm levels in *SYNM* in relation to PTSD-related conditions (e.g., T2D) and molecular mechanisms, such as mitochondria dysfunction.

In addition to differential analysis at CpG sites, our analyses revealed thirteen DMRs (FDR < 0.05) associated with PTSD, confirming some prior findings, and highlighting novel hits for further validation. Among the DMRs identified, chr21:45705428-45706044, mapped to a transcription start site in *AIRE*, an autoimmune regulator gene. *AIRE* is highly expressed in both central and peripheral organs and plays an important role in immunity by regulating autoantigens and autoreactive T-cell activity^85,86^. The *AIRE* promoter is hypermethylated in the absence of any inflammatory stimuli^87^ and the role of inflammation in PTSD etiology is well-established^88^. Moreover, a DMR in *AIRE* has been previously associated with childhood physical neglect^89^; a sub-type of childhood trauma, which is a known risk factor for developing PTSD^90,91^. Differential *AIRE* methylation seen here could, through a biological pathway (e.g., inflammation), link underlying risk factors (e.g., childhood trauma) involved in susceptibility to developing PTSD. This plausible, but putative mechanistic pathway between childhood trauma and PTSD requires further validation and exploration. Another PTSD-associated DMR mapped to the brain-derived neurotrophic factor antisense (*BDNF-AS*) – a lincRNA shown to regulate *BDNF* expression, which is important for neuronal differentiation, learning and memory^92^. Our findings align with previous EWAS that reported differential *BDNF* methylation in individuals who experienced childhood trauma^93,94^ and those with PTSD^95^.

PTSD and MetS are known risk factors for CVDs and mortality^20,21^, and a role for accelerated cellular aging in PTSD-related CVD risk has been proposed^96–98^. Here, we provide evidence for accelerated DNAm age for PTSD-MetS comorbidity across three DNAm age predictors — Horvath, Hannum, and Levine PhenoAge. Our findings align with a recent study showing substantially higher likelihood of accelerated GrimAge among U.S veterans with PTSD and obesity^43^, and suggesting a synergistic effect between PTSD-related CVD risk and accelerated biological aging. We could not replicate previous associations between PTSD diagnosis and accelerated aging based on Hannum and Horvath clocks in European-Americans^29,99,100^ or GrimAge in African-Americans^39^. Rather, we found an association between PTSD severity and the Horvath DNAm age clock. This disparity could be due to the improved statistical power of using quantitative measures of PTSD severity, which capture disease variation better than binary case-control measures. Differences may also stem from variations in diagnostic tools (self-report vs. clinical diagnosis) or age differences between cohorts. Biological causal pathways could also explain the link between PTSD severity and accelerated aging. Notably, the Horvath’s DNAm age clock is enriched for glucocorticoid response elements linked to stress^101^. It is plausible that chronic stress responses experienced by individuals with severe PTSD can affect the body’s cellular processes and lead to epigenetic changes associated with aging. Dysregulated inflammatory responses are one potential type of cellular process involved in this association^39,40,102^. Whether the severity of PTSD directly causes immune-related dysregulation, or whether biological aging is a readout of a systemic vulnerability to develop PTSD and other age-related health conditions warrants further research.

The findings from this study should be interpreted within the context of several limitations. Though our study benefits from having a deeply-phenotyped and genetically admixed population, the sample size (N = 120) lacks sufficient power to detect significant epigenome-wide effect sizes similar to those reported in African Americans by Katrinli et al. (N = 855)^39^. Moreover, given the small sample size, we could not examine the potential interaction of MetS with PTSD in the EWAS, as this would require dividing the sample into sub-groups with and without MetS for the case and control groups. Second, DNAm was profiled using whole blood as a surrogate for the brain. Though previous research supports a blood-brain concordance in DNAm levels for PTSD^103^, some DNAm sites show inconsistent patterns in the brain and blood^104^. Therefore, caution is needed when inferring DNAm levels across these tissues. Third, we could not account for the role of other confounders (such as diet, exercise, depressive symptoms, and CUD), which may impact the associations reported in the EWAS and DMR analyses.

## Data Availability

All data produced in the present study are available upon reasonable request to the authors

## Acknowledgements

This research is supported by: (i) The South African Research Chair in PTSD hosted by Stellenbosch University, funded by the Department of Science and Technology (DST) and administered by the South African National Research Foundation (NRF) and (ii) South African Medical Research Council in terms of the SAMRC’s Flagships Awards Project (Grant no: MRC-RFA-IFSP-01-2013/SHARED ROOTS). The work by Leigh van den Heuvel was supported in part by the National Research Foundation of South Africa (Grant Number 138430) and by the South African Medical Research Council under a Self-Initiated Research Grant. The content hereof is the sole responsibility of the authors and do not necessarily represent the official views of the SAMRC or the funders.

## Data Availability

Available upon reasonable request.

## Code Availability

The scripts used to perform the EWAS are available via PGC-PTSD EWAS GitHub (https://github.com/PGC-PTSD-EWAS/). DMR analysis was performed using dmrff (https://github.com/perishky/dmrff). DNAm age analysis was conducted using dnaMethyAge (https://github.com/yiluyucheng/dnaMethyAge).

---

**Supplementary Figure 1:**
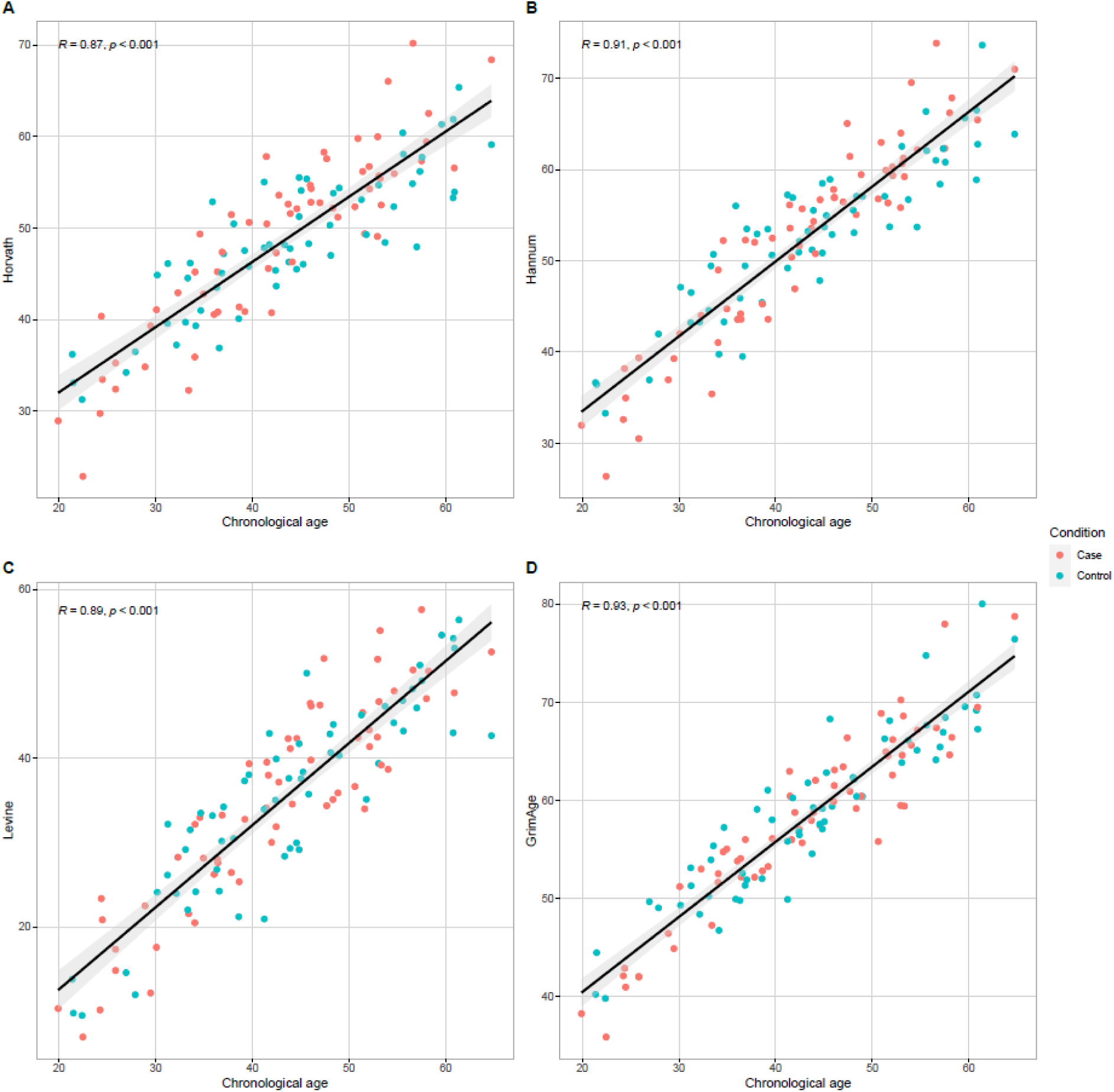
Correlation estimates for all four DNAm age clocks. Epigenetic age across all DNAm clocks was significantly positively correlated (*p* < 0.001).

**Supplementary Figure 2:**
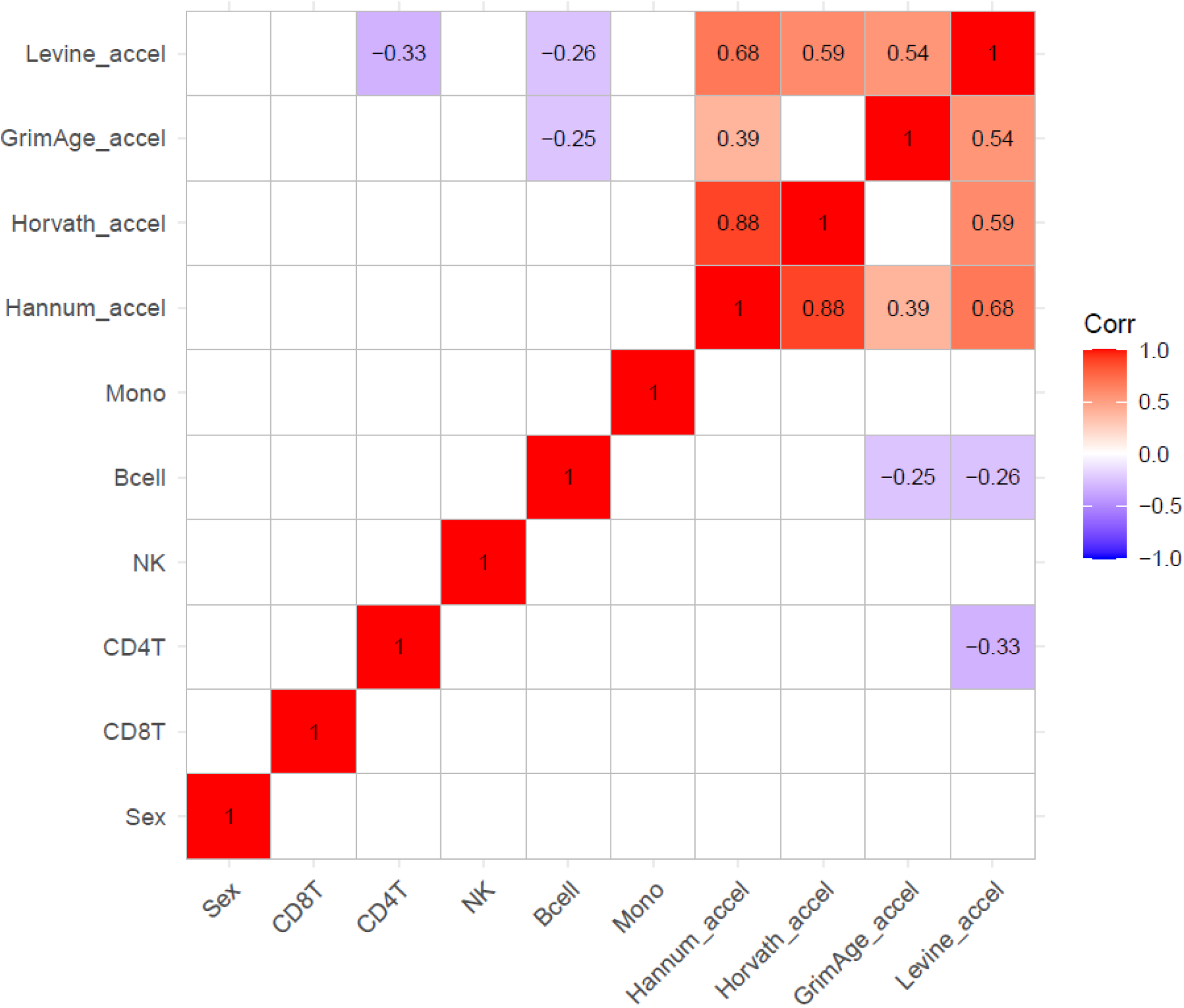
Correlation estimates for epigenetic age acceleration and study covariates. Epigenetic age acceleration was negatively correlated with B cells (GrimAge: r = - 0.25; *p* = 0.04; Levine PhenoAge: r = -0.26; *p* = 0.04) and CD4 T cells (Levine PhenoAge: r = - 0.33; *p* = 0.01).

